# Depression, Anxiety and Stress among the Patient of Chronic Kidney Disease at Nadiad city, A Cross sectional survey

**DOI:** 10.1101/2021.08.01.21261443

**Authors:** Kailash Nagar, Arpita Vaidya, Khushabu Patel

## Abstract

**Introduction:** Chronic kidney disease is considered as a public health problem worldwide [1]. It is defined by kidney tissue injury with or without a decrease in glomerular filtration rate and /or a decrease in kidney function over a period of three or more months. When the glomerular filtration rate [GFR] is below 15ml./min./1.73m^2^, the patient is in the terminal stage or dialysis, requiring renal replacement therapy [RRT], dialysis or transplant as alternative treatments. Renal failure is the inability of the kidney to excrete wastes, concentrate urine, and conserve electrolytes. Renal failure is precipitated from a variety of etiological factors. It is treatable but not curable, which means that the patient needs a long term therapies or transplantation [2].

The overall prevalence of depression, anxiety and stress in this study found to be 22.9%, 19.2%, and 28.2%, respectively. Forty nine (13.84%) respondents had mild depression twenty eight (7.91%) has moderate depression[2].

Depression and anxiety are frequent comorbid disorders among chronic kidney disease (CKD) patients, with estimated prevalence of approximately 25% in this population, [3] and are associated with worse outcomes, such as progression to end-stage renal disease (ESRD) and mortality [4,5,6,7]. The transition from predialysis management to renal replacement therapy (RRT) is a stressful event in the course of CKD, leading to challenges and decisions that might increase their susceptibility to anxiety, mood disorders or even exacerbate psychological issues that already exist [8].

**Aims:** The researcher aim to assess the level of depression, anxiety and stress among chronic kidney disease patient in selected hospital of Nadiad City, Gujarat. The research wants to explore the actual psychological status of the patients who were suffering from chronic illness like CKD, because due to long term suffering from the illness disturbed the everyone mind and cause various psychological changes in the body. Hence research want to assess what kind of changes take place due to chronic kidney diseases.

**Objectives:**

1. To assess the level of depression among CKD patients at selected hospital, Nadiad.
2. To assess the level of anxiety among CKD patients at selected hospital, Nadiad.
3. To assess the level of stress among the CKD patients at selected hospital, Nadiad.

**Methodology:** *Design and Setting:* Descriptive cross sectional survey research design was adopted for the study and non-probability purposive sampling method was used to drawn samples from the participants. For the data collection researcher has used modified The DASS-42 is a 42 item self-report scale designed to measure the negative emotional states of depression, anxiety and stress, it is a standardized tool. Prior to data collection written setting permission obtain from the head of the hospital as well prior inform consent form was obtain from the study participants, the objectives and methods of the study were appropriately explain to the samples. For data collection researcher has select MPUH kidney hospital situated Nadiad City. The total sample size was 30 chronic kidney disease patients. The research data collection tool consist of following Section I Demographic variables of the CKD patients section II DASS 42 questionnaire self rating scale.

*Statistical Analysis:* Descriptive statistics applied where, data were analyzed by using SPSS software, and Frequency, percentage, tables etc. were used to represent the statistical data in the tables and graph and figure.

*Result:* The majority of participants (30%) were 41-50 years, sample (36.67%) belong age group of above 50 years, majority 56.67% were Male, (43.33%) were graduate, (67%) Having 5000-10000 Monthly Income, (60%) were living in Joint family, (56.67%) were belong to 0-3 years, (23.33%) were belong above 9 years if illness. Duration of hospitalization (80%) 0-15 days. Prevalence rate of depression, anxiety and stress among Chronic Kidney Disease the most of patient have 50%, had moderate symptoms of depression, anxiety and stress, 30% had mild and only 20% have severe symptoms of depression, anxiety and stress which was measured by DASS self rating scale.

*Conclusions:* The currents study ended to assess the prevalence rate of the depression, anxiety and stress among Chronic Kidney Disease patients, the study result concluded that the majority (50%) of Patients having moderate level of depression, anxiety and stress. The people in age group 41-50 or above 50 are having higher rate of depression, anxiety, and stress during chronic kidney disease.

## INTRODUCTION

Chronic kidney disease is considered as a public health problem worldwide [1]. It is defined by kidney tissue injury [with or without a decrease in glomerular filtration rate and /or a decrease in kidney function over a period of three or more months. When the glomerular filtration rate [GFR] is below 15ml./min./1.73m^2^, the patient is in the terminal stage or dialysis, requiring renal replacement therapy [RRT], dialysis or transplant as alternative treatments. Renal failure is the inability of the kidney to excrete wastes, concentrate urine, and conserve electrolytes. Renal failure is precipitated from a variety of etiological factors. It is treatable but not curable, which means that the patient needs a long term therapies or transplantation [2].

The overall prevalence of depression, anxiety and stress in this study found to be 22.9%, 19.2%, and 28.2%, respectively. Forty nine (13.84%) respondents had mild depression twenty eight (7.91%) has moderate depression [2].

The patients with renal disease suffer higher rates of depression including their caregivers who also suffer from mental stress[9,10].

Depression is recognized as the most common psychiatric problem in patients with end-stage renal disease. Stress negatively affects the quality of life of not only the patients on hemodialysis but also their caregivers [10].

Mood disorders, including anxiety and depression, are prevalent among patients with chronic kidney disease (CKD) who are on hemodialysis. Anxiety and\or depressive symptoms among those patients have been associated with early initiation of dialysis and adverse outcome [11].

The aim of this study is to determine the prevalence of anxiety and depression among CKD patients undergoing hemodialysis. Thus, the findings of this study will serve as a basis to initiate a needs assessment among CKD patients experiencing anxiety and depression and to develop and implement a support management plan to improve mental health services for these CKD patients and improve their quality of life[11]

## OBJECTIVES

### Assumption

The study is based on following assumption:

1. The patients with CKD may have higher level of depression anxiety and stress.

### Operational Definitions

1. **Depression:-** It refers to abnormal extension of over elaboration of sadness and grief and loss of interest in pleasurable activities feeling of worthlessness and excessive guilt among CKD patients as measured by DASS self – rating scale.
2. **Anxiety:-** It refers to diffuse apprehension that is vague in nature and is associated with feeling of uncertainly by DASS self-rating scale.
3. **Stress**:- It refers to a stage of psychological and physiological imbalance from the disparity between situational demand and individual ability measured by DASS Self rating scale.
4. **CKD**:- It refers to a chronic condition in which patients kidney is no longer able to perform its function.

### Delimitation:-

A study is delimited:-

1. The study was limited to the patients with CKD at selected hospital, Nadiad.
2. The sample size was limited to 30 CKD patients.

## METHODOLOGY

### Research Approach :-

Non-experimental research approach

### Reseach Design :-

cross-sectional study research approach was used

### Variable:-

#### Background variable includes

Age, gender, education, economic status, family type, Duration of Disease condition, Duration of Hospitalization.

#### Dependent variable

Depression, Stress and Anxiety.

#### Independent variable

DASS self rating scale

#### Population of The Study

The population includes all the patient of CKD admitted in MPUH hospital of the Nadiad city.

#### Sample and Sampling Techniques

The sample size was 30 CKD patients and the sampling techniques was non-probability convenient sampling techniques.

#### Tool for Data Collection

DASS self rating scale was used to Assess the level of Depression, Stress and Anxiety.

## RESULT & FINDINGS

**[Table/Fig-1]** Revealed that the distribution of sample according to age sample 6 (20%) belong age group of 20-30 years, sample 4(13.33%) belong age group of 31-40 years, sample 9(30%) belong age group of 41-50 years, sample 11(36.67%) belong age group of above 50 years.

**[Table/Fig-1].** Frequency and Percentage distribution of CKD Patients according to demographic variables.

| Variables | Categories | (F) | (%) |
| --- | --- | --- | --- |
| Age | 20-30 years | 6 | 20 |
|  | 31-40 years | 4 | 13.33 |
|  | 41-50 years | 9 | 30 |
|  | Above 50 years | 11 | 36.67 |
| Gender | Female | 13 | 43.33 |
|  | Male | 17 | 56.67 |
| Education | Illiterate | 4 | 13.33 |
|  | Primary | 3 | 10 |
|  | Secondary | 3 | 10 |
|  | Higher secondary | 7 | 23.3 |
|  | Graduate or Above | 13 | 43.33 |
| Monthly Income | 5000-10000 | 11 | 36.67 |
|  | 10000-15000 | 6 | 20 |
|  | 15000-20000 | 3 | 10 |
|  | above 20000 | 10 | 33.33 |
| Family Type | Joint | 18 | 60 |
|  | Nuclear | 8 | 26.67 |
|  | Single | 4 | 13.33 |
| Duration of disease condition | 0-3 years | 17 | 56.67 |
|  | 4-6 years | 2 | 6.67 |
|  | 7-9 years | 4 | 13.33 |
|  | > 9 years | 7 | 23.33 |
| Duration of hospitalization | 0-15 days | 24 | 80 |
|  | 16-30 days | 2 | 6.67 |
|  | 31-45 days | 0 | 0 |
|  | > 45 days | 4 | 13.33 |

Regarding the Gender CKD patients out of 30 samples, 13(43.33%) were Female and 17(56.67%) were Male.

Regarding the Education CKD patients out of 30 samples, 4(13.33%) were illiterate, 3(10%) were primary education, 3(10%) were secondary education, 7(23.33%) were higher secondary education, 13(43.33%) were graduate or above.

Regarding the Economical condition of CKD patients out of 30 samples, 11(67%) Having 5000-10000 Monthly Income, 6(20%) Having 10000-15000, 3(10%) having 15000-20000, 10(33.33%) having above 20000 rupees monthly.

Regarding the Type of family patients of CKD out of 30 Samples, 18(60%) were living in Joint family, 8(26.67%) were living in Nuclear family, 4(13.33%) were living single.

Regarding the Duration of Disease Condition of CKD patients out of 30 samples, 17(56.67%) were belong to 0-3 years, 2(6.67%) were belong to 4-6 years, 4(13.33%) were belong to 7-9 years, 7(23.33%) were belong above 9 years.

Regarding to Duration of Hospitalization of patients out of 30 samples, 24(80%) were belong 0-15 days, 2(6.67%) were belong to 16-30 days, 0(0%) were belong to 31-45 days, 04(13.33%) were belong to above 45 Days.

Regarding to participation in any research activity related CKD out of 30 samples, 04(13.33%) has been participated and 26(86.67%) has been no participated.

**[Table/Fig-2]** Revealed that the distribution of sample according to incidence rate of Depression, Anxiety and Stress, 9(30%) were suffering from mild symptoms of Depression, Anxiety and Stress, 15(50%) were suffering from moderate symptoms of Depression, Anxiety and Stress, 6(20%) were suffering from severe symptoms of Depression, Anxiety and Stress.

**[Table/Fig-2].** Frequency and percentage distribution of CKD Patients according to the Incidence rate of Depression, Anxiety and Stress.

| Score | Number of Sample | Percentage (%) |
| --- | --- | --- |
| Mild | 9 | 30% |
| Moderate | 15 | 50% |
| Severe | 6 | 20% |
| **Total** | **30** | **100%** |

**[Table/Fig-3].**
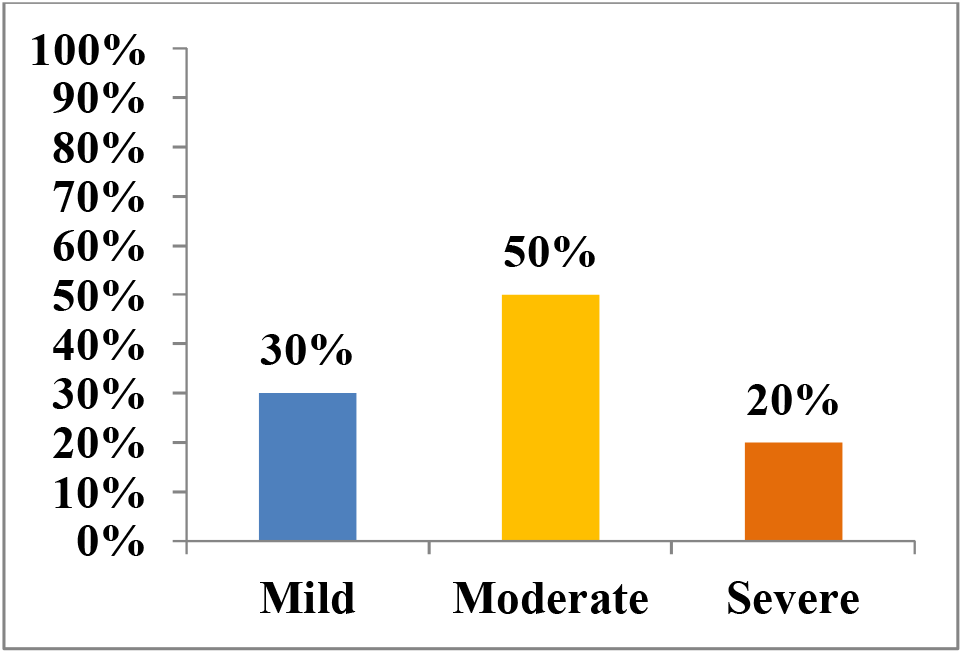
Bar diagram Percentage distribution of CKD Patients according to the Incidence rate of Depression, Anxiety and Stress.

### Major Finding of Study

Finding related to distribution of sample according to age sample 6 (20%) belong age group of 20-30 years, sample 4(13.33%) belong age group of 31-40 years, sample 9(30%) belong age group of 41-50 years, sample 11(36.67%) belong age group of above 50 years

Finding related to distribution of sample according to incidence rate of Depression, Anxiety and Stress, 9(30%) were suffering from mild symptoms of Depression, Anxiety and Stress, 15(50%) were suffering from moderate symptoms of Depression, Anxiety and Stress, 6(20%) were suffering from severe symptoms of Depression, Anxiety and Stress.

## Conclusion

The currents study ended to assess the prevalence rate of the depression, anxiety and stress among Chronic Kidney Disease patients, the study result concluded that the majority (50%) of Patients having moderate level of depression, anxiety and stress. The people in age group 41-50 or above 50 are having higher rate of depression, anxiety, and stress during chronic kidney disease.

## Data Availability

as per the guideline of the manuscript submission all the data has been included

## Ethical Clearance

The study was approved by the research committee, IEC – DPCN/1^st^ IEC/2018-19/09 and a formal written permission was gathered from the authority of Principal of Institute.

## Statement of Informed consent

Informed consent was acquired from the participants prior to data collection.

